# Mathematical Modeling & the Transmission Dynamics of SARS-CoV-2 in Cali, Colombia: *Implications to a 2020 Outbreak & public health preparedness*

**DOI:** 10.1101/2020.05.06.20093526

**Authors:** Jorge Humberto Rojas, Marlio Paredes, Malay Banerjee, Olcay Akman, Anuj Mubayi

## Abstract

**Introduction:** As SARS-COV-2 and the disease COVID-19 is sweeping through countries after countries around the globe, it is critical to understand potential burden of a future outbreak in cities of Colombia. This pandemic has affected most of the countries in the world because the high global movement of individuals and excessive cost in interventions.

**Objective:** Using demographic data from city of Cali, disease epidemiological information from affected countries and mathematical models, we estimated the rate of initial exponential growth of new cases and the basic reproductive rate for a potential outbreak in city of Cali in Colombia.

**Materials and methods:** We used dynamical models with different modeling assumptions such as use of various types of interventions and/or epidemiological characteristics to compare and contrast the differences between Colombian cities and between Latin American countries.

**Results:** Under the assumption of homogeneously mixing population and limited resources, we predicted expected number of infected, hospitalized, in Intensive Care Units (ICU) and deaths during this potential COVID-19 outbreak. Our results suggest that on a given day in Cali there may be up to around 73000 cases who might need hospitalization under no intervention. However, this number drastically reduces if we carry out only-isolation intervention (with 16 days of symptomatic infection; ~13,000 cases) versus both quarantining for 6 days and isolation within 16 days (~3500 cases). The peak in Cali will reach in 2-3 months.

**Conclusions:** The estimates from these studies provides different scenarios of outbreaks and can help Cali to be better prepared during the ongoing COVID-19 outbreak.

## 1. Introduction

An outbreak of pneumonia with unknown etiology in the Wuhan city, Hubei’s province in Mainland China was first reported on December 31, 2019 by World Health Organization (WHO). On January 7, 2020 a virus associated with this outbreak was isolated [1] and found to be novel strain of Coronavirus that may have crossed over to human population from its sylvatic host. On January 30, 2020, WHO declared the Public Health emergency of International importance [2] and denominated this coronavirus as SARS-COV-2 while the disease was referred as COVID-19 on February 12, 2020 [3]. This virus belongs to the genus betacoronavirus of the subfamily coronavirus and family Coronaviridae. 2002-2004 SARS epidemic and 2012-2016 MERS epidemic were also caused by the viruses belonging to the family coronavirus, however, both showed lower transmissibility and absolute lethality [4].

In order to protect people from this unknown outbreak of COVID-19 it is critical to understand its trends and estimate potential burden. Mathematical models have been used before to this purpose for various epidemic diseases and can be extremely helpful in disease preparedness. An important threshold quantity associated with a disease transmissibility is the basic reproduction number, denoted by *R*_0_ (pronounced “R naught”). The epidemiological definition of *R*_0_ is the average number of new cases of the disease that will be generated by one contagious person during his/her infectious period. It specifically applies to a population of people who were previously free of infection and not vaccinated. Three possibilities exist for the potential spread or decline of a disease, depending on its *R*_0_ value: (i). If *R*_0_ is less than 1, each existing infection causes less than 1 new infection. In this case, the disease will decline and eventually disappear. (ii). If *R*_0_ equals 1, the disease will stay alive, but there won’t be an epidemic. (iii). If *R*_0_ is greater than 1, cases could grow exponentially and cause an epidemic or even a pandemic [5]. A preliminary *R*_0_ estimate of 1.4 - 2.5 was presented on Jan 23, 2020 in a WHO’s statement regarding the outbreak of 2019-nCoV [1]. S. Zhao et al. [6] estimated the mean *R*_0_ for 2019-nCoV in the early phase of the outbreak ranging from 3.3 to 5.5 (likely to be below 5 but above 3 with rising report rate) [6,7], which appeared slightly higher than those of SARS-CoV (*R*_0_:2−5) [4]. J. Read et al. [44] estimated the *R*_0_ for 2019-nCoV to be in the range 3.6 - 4.0, indicating that 72-75% of transmissions must be prevented in order to stop the increasing trend. The explosive and dramatic behavior of the spread of this virus SARS-CoV-2 in mainland China, has forced health and administrative authorities take severe and strict control measures. Until January 31, 2020, 11950 cases had been reported and 259 deaths but the number of cases had increased to 75,184 and 2009 deaths on February 18, 2020 [8]. The pharmacological treatment with antivirals is unknown and the vaccine is in the early stages of investigation.

Cali is a city with a population of about 2.5 million (2,492,442 people, 98.4% in urban areas and 1.6% in rural areas) [9], located at 3°27’26’’ of latitude North and 76°31’42’’ of longitude West (Greenwich Meridian), at an altitude of 1,070 meters above the sea level, with an average temperature of 24.7°C, an annual precipitation of 1,019.2 mm 12. Cali presents two rainy seasons during the year [10], the first one between the months March and May, and then the second one rainy period occurs between September and November when the temperatures come down. Other strains of Coronavirus are known to be significantly related to climatic conditions with colder weather more suitable for their spread. It is worth considering that the covid-19 epidemic in Mainland China began in winter, the period of year with lower temperature and that in Cali the first winter season is approaching now in April/May when the temperature drops, and the population tends to be more crowded. Cali is a touristic city and the number of passengers moved by international routes in Colombia [11] reached 11,527,416 people alone in the first half-year. The countries of origin of these flights were of North America with 3,108,146 people, Europe 4,024,519 people, South America 2,608,066 people. Between the factors [12] explain the emergence or re-emergence of infectious diseases the modern aeronautical technology allows the connection between countries in very short time and thus sick people carry infectious agents over long distances [13]. This information will allow prepare plans of education to community that interrupts the transmission of the virus.

*The objectives of this study are to (i) use available current COVID-19 information from other countries and estimate, for the city of Cali, Colombia, the expected number of cases that needs to be hospitalized over an outbreak, maximum daily number of cases that can be expected, time needed to reach the epidemic peak and mean duration of an epidemic for the current outbreak in the presence of different control actions by the public health department and different modeling assumptions, (ii) compare situation in Cali with other four major cities of Colombia, and (iii) compare the situation in Colombia with other similar countries.*

## 2. Materials and Methods

We considered different modeling assumptions to capture COVID-19 outbreak in Cali and four other cities of Colombia. In order to understand the ongoing burden of the disease, a systematic procedure was implemented. The procedure in this study primarily involves four steps: (i) Using reported estimates of *R*_0_ and model-derived formulation of *R*_0_ (corresponding to different model and under the assumption of initial exponential growth rate of epidemic), estimate effective transmission rate in the population, (ii) Use estimated effective transmission rate, demographic data from city of Cali and epidemic models, to predict different outbreak scenarios for Cali, and (iii) From these outbreak scenarios (in step (ii)), estimate the total number of cases, hospitalizations, time needed to reach the peak and expected duration of the outbreak.

### Basic Modeling Framework

#### Mathematical Model to Capture Potential Epidemic in Cali, Colombia

For the mathematical modeling of the transmission dynamics of the new SARS-CoV-2 coronavirus, we have established the structure compartmental of SEIR model (Susceptible, Exposed, Infectious, Recovered) and applying the method of differential equations 5 based on the COVID-19 epidemic data in continental China [14,15].

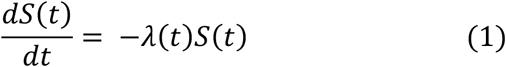

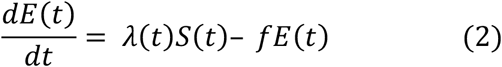

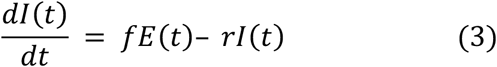

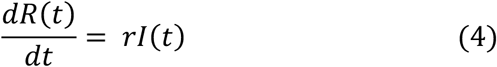

Model variables and parameters are defined in Table 1. The rest of the models are defined in Table 2 and in the Appendix, section B.

**Table 1.** Variables and parameters of the SEIR model

|  | Symbol | Definition |
| --- | --- | --- |
| Variables | $S$ | Susceptible population |
| | $E$ | Exposed (pre-infectious) population |
| | $I$ | Infectious population |
| | $R$ | Recovered population |
| Parameters | $\lambda$ | Rate at which susceptible individuals becoming infected per unit time, at time $t$ |
| | $f$ | Rate at which individuals in the pre-infectious category become infectious per unit time |
| | $r$ | Rate at which infectious individuals recover per unit time |

**Table 2.** Formulas to the estimate basic reproduction number

| $R_0/R_c$ in terms of $\Lambda$ | Assumptions | Model (Actual $R_0$) |
| --- | --- | --- |
| $1 + \Lambda D$ | This expression assumes that the pre-infectious period is very short in comparison with the infectious period, or individuals are infectious immediately after they are infected. The infectious period is assumed to follow the exponential distribution. (Appendix, section B.1) | SIR<br>($R_0 = \frac{\beta N}{\alpha}$) |
| $1 + \Lambda T_s$ | This expression is typically used when the pre-infectious and/or infectious periods are not known but are assumed to follow the exponential distribution. It can be shown that if the pre-infectious and infectious periods follow the exponential distribution, then the serial interval ($T_s$) equals $D + D'$. (Appendix, section B.2) | SEIR with only I Infectious<br>($R_0 = \frac{\beta N}{\alpha}$) |
| $1 + \left(\frac{\ln 2}{T_d}\right) D$ | This equation can be derived from the first one in this table using the fact that the time until the number of infectious persons doubles relative to that at some other time ($T_d$) is related to the growth rate through the equation $\Lambda = \frac{\ln 2}{T_d}$. (Appendix, section B.2) | |
| $1 + \Lambda T_s + f_r(1 - f_r)(\Lambda T_s)^2$ | This expression is a more detailed version of the third one in this table and is typically used when the pre-infectious and/or infectious periods are poorly understood but are assumed to follow the exponential distributions. $f_r = \frac{D}{T_s}$ is the ratio between the infectious period and the serial interval. (Appendix, section B.2) | |
| $(1 + \Lambda D)(1 + \Lambda D')$ | The pre-infectious and infectious periods follow the exponential distribution. Neither the pre-infectious nor infectious periods are short, relative to the other. (Appendix, section B.2) | |
| $\frac{(1 + \Lambda D')(1 + \Lambda D) \left( 1 + \frac{1}{D D' \varepsilon} \right)}{\left( 1 + \Lambda D + \frac{1}{D D' \varepsilon} \right)}$ | It is assumed that E individuals are infectious and transmit infection, however, their infectivity is lower than that of I individuals. (Appendix, section B.3) | SEIR with E & I infectious<br>$(R_0 = \frac{\beta N}{\alpha} + \frac{\varepsilon \beta N}{\kappa})$ |
| $\frac{(\eta + \delta \gamma)(\Lambda + \eta)(\Lambda + \alpha + \gamma)}{\eta(\alpha + \gamma)(\Lambda + \eta + \delta \gamma)}$ | It assumes that there is no E stage (i.e., no asymptomatic stage) but there are some therapeutic effects on patients in isolation. (Appendix, section B.4) | SITR, model with treatment<br>$(R_0 = \frac{\beta N}{\alpha + \gamma} \left( 1 + \frac{\delta \gamma}{\eta} \right))$ |
| $\frac{(\sigma + \alpha_1 + \frac{\gamma_1}{q})(\Lambda + \sigma + \alpha_1)(\Lambda + \gamma_1 + \phi)}{(\gamma_1 + \phi)(\sigma + \alpha_1)(\Lambda + \sigma + \alpha_1 + \frac{\gamma_1}{q})}$ | It assumes SEIR type model but with quarantining for suspected cases and isolation for confirmed cases. Due to these measures (Appendix, section B.6) | SEIR-Q<sub>1</sub>Q<sub>2</sub>Q<sub>3</sub> Model with standard incidence<br>$(R_c = \frac{\beta q}{\gamma_1 + \phi} + \frac{\beta \gamma_1}{(\gamma_1 + \phi)(\sigma + \alpha_1)})$ |
| $R_c(\Lambda)$ which is given in appendix | It assumes that hospitalization of individuals is possible quickly and asymptomatic/symptomatic both are infectious. (Appendix, section B.7) | SEIJR Model with standard incidence<br>$(R_c = \frac{\beta q}{\eta + \kappa + \mu} + \frac{\beta \kappa}{(\eta + \kappa + \mu)(\gamma_1 + \alpha + \delta)} + \frac{\beta I(\alpha \eta + \alpha \kappa + \eta \delta + \gamma_1 \eta)}{(\eta + \kappa + \mu)(\gamma_1 + \alpha + \delta)(\gamma_2 + \delta)})$ |

#### Model-related metrics

Consider a simple epidemic model, Susceptible-Infectious-Recovered (SIR) or Susceptible-Exposed-Infectious-Recovered (SEIR) model with I individuals are only infectious. The number of susceptible individuals who are infected per unit time is given by the product of the force of infection, *λ*(*t*), and the number of susceptible individuals at time *t* (*i. e., λ*(*t*)*S*(*t*)) under the assumption that individuals contact each other randomly. Let *λ*(*t*) is the force of infection or rate at which a susceptible individual is infected or rate at which they move to an infected compartment then it can be written as:

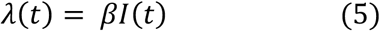

where *β* is the rate at which two specific individuals come into effective contact per unit time and *I*(*t*) is the number of infectious individuals at time *t*. The formula for basic reproduction number of the model is *R*_0_ *= βND* and can used it to estimate effective contact rate as

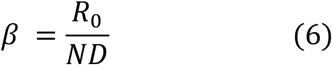

where *N* is the population size at the beginning of an outbreak and *D* is the length of the infectious period.

Now if Exposed state (defined as individuals who are infected not infectious) is present in the model then the model will be referred as SEIR model. Suppose *f* is the rate at which the individuals move from exposed compartment to infectious compartment. The rate *f =* 1*/D′*, where *D′* is the duration of the latent or pre infectious period. Let *r* be the rate at which the individuals move from infectious compartment to recovered or immune compartment, i.e., *r =* 1*/D*, where *D* is the duration of the infectious period.

Since *R*_0_ is a metric computed early in the epidemic, it can be used to relate it with the initial growth rate of an epidemic. Different formulae are used to estimate *R*_0_ using data from the early stages of an epidemic, each of them requires estimates of something which is often referred to as exponential growth of rate of the epidemic sometimes denoted by Λ (capital lambda). During the early stages of an epidemic, the number of infectious individuals increases at an approximately constant exponential growth rate.

### Estimation of Various Relevant Quantities

#### Initial Exponential Growth Rate

We can estimate this growth rate as follows. Consider an exponential growing function of number of reported cases *I*(*t*) where Λ is the exponential growth rate parameter, i.e.,

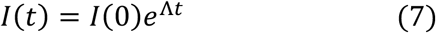

where *I*(0) is the initial number of infectious individuals at the beginning of infection. If we take natural logarithm of this equation, we obtain the following equation

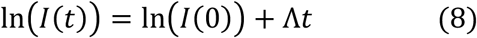

relating the number of infectious individuals and Λ. Note, this is the equation of a straight line with slope Λ, suggesting that if we plot the natural logarithm of the number of infectious individuals against time, we should obtain a straight line with slope Λ.

On the other hand, we can also write an expression of *R*_0_, in terms of Λ. This expression of *R*_0_ will depend on the type of model considered and other model parameters including *D’* and *D* the average durations of the pre-infectious and infectious periods respectively. Sometimes *R*_0_ can also be expressed in terms of the serial interval, *T_s_* (defined as the time between the start of symptoms in the primary patient (infector) and onset of symptoms in the patient receiving that infection from the infector (the infected) [7]. In Table 2 the different formulas are shown. For example, in case of the SEIR model with only *I* infectious, the basic reproduction number can be written as

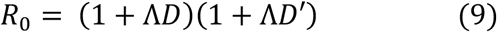

and in case of the SIR model, *R*_0_ *=* 1 *+* Λ*D*.

Furthermore, we can use equation (8) to obtain the relationship between the growth rate Λ and the doubling time *T_d_* of an epidemic, which is defined as the time until the number of cases in the population doubles, relative to that at some other time. Suppose that there is only one infectious individual at time *t =* 0 (*I*(0) *=* 1) and there are two infectious individuals at time *t = T_d_* (*I*(*T_d_*) *=* 2). Substituting for *I*(0) *=* 1 and *I*(*T_d_*) *=* 2 into equation (8) we obtain *ln*2 *= ln*1 *+* Λ*T_d_* which implies

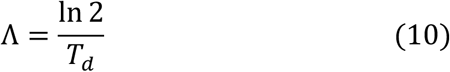

Since R_0_ can be written in the form of Λ and the doubling time can be computed as in equation (10), we can find *T_d_* in terms of *R*_0_. For example, in case of SIR model,

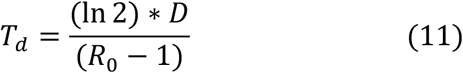

and in case of SEIR model with only *I* infectious, it can be written as

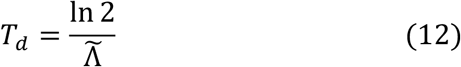

where 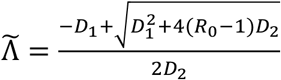, *D*_1_*= D + D′* and *D_2_ = D * D′*. This formula can be used to study the role of epidemiological factors such as transmission rate (*β*), asymptomatic and symptomatic periods.

#### Estimation of Model Parameters

In groups of Japanese migrants who were repatriated, the proportion of positive asymptomatics to PCR test, was estimated by Hiroshi Nishiura et al, in 41.6% (CI 95%: 16.7-66.7) [16]; Anne Kimball et al, [17] found 57.0% and Kenji Mizumoto et al 51.7% [18]. For purpose of our study we will take 50 % as parameter value [19]. It has also been established that 14% of the symptomatic people are hospitalized for complications related to pneumonia and respiratory distress [20,21]. From those hospitalized, 15% die [22-24]. Because the covid-19 is novel disease, some parameters are unknown or are known with less precision, and hence, they are estimated. Using case reports, outbreaks’ studies and some based on the behavior of coronavirus in past epidemics, these parameters are estimated. Multiple research articles report incubation period estimates obtained from different methods. The most of them report incubation period mean between 5 – 7 days and the range between 2 – 14 days [25-34]. We have used the incubation period to be 6.4 days based on most of the studies. Although the virus SARS-2 can be detected in nasopharyngeal swab between 2.5 days before and 18 days after starting symptoms, infectivity is considered very low after 7 days. The infectious period is assumed for this study, an average of 7.6 days based on reported 14 days quarantine period COVID-19 [35-39].

#### Estimation of R_0_

The data used for calculating *R*_0_ were collected from published research in the New England journal of Medicine on January 29, 2020 [14]. The novel coronavirus SARS-CoV-2, causes COVID-19, which is characterized mainly by fever, muscle pain and cough and it can move along to severe phases of pneumonia or even die. Because it is a new disease, there is a small knowledge about the time of latent period, infectious period in the natural history of disease, though some estimates have been made based on cluster of cases and hospitalized.

### Models

In this study, we consider various modeling assumptions leading to seven different models, six of them are defined in Table 2 and their details are given in Appendix B. The last (seventh) model is referred as Age of infection model, whose some information are given below and other details are given in Appendix B. Suppose *S,E,I,R,T,J, Q*_1_*, Q*_2_ and *Q*_3_ represent Susceptible, Exposed, Infectious, Recovered, Treated, Hospitalized, Susceptible-Quarantined, Exposed-Quarantined and Infectious-Isolated. The models are (i) simple SIR, (ii) simple SEIR with I only infectious, (iii) simple SEIR with E and I infectious, (iv) SITR, (v) SIR with general distribution of age of infection (vi) SEIR-Q_1_Q_2_Q_3_, and (vii) SEIJR.

#### Age of Infection Model

*S*(*t*) and *φ*(*t*) denote the number of susceptibles and total infectivity at time *t*. The total infectivity is measured as the average of product of the number of infected individuals and the mean infectivity of the infected individuals at their age of infection *τ* where *τ* ∈ [0,*t*]. Denoting the mean infectivity at time *τ* by *A*(*τ*), we can write the epidemic model with age of infection as follows

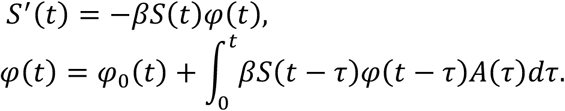

Then the basic reproduction number is defined as

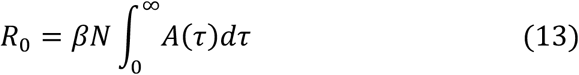

The basic reproduction number in terms of initial growth rate is as follows,

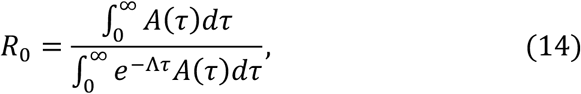

where Λ is a positive eigenvalue satisfying the relation

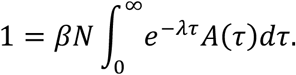

#### Interventions

The modeling assumptions in the models were related to epidemiological characteristics and/or type of interventions. The interventions that we have considered include quarantine, isolation, treatment, or infectious hospitalized individuals with mild infection.

We also consider three different types of interventions [40] such as

- **“Wuhan-style” Containment:** It assumes closed borders with full shutdown and no movement of individuals (first reporting Jan 1 in Wuhan and this type of lockdown started on Jan 23). In order to capture this intervention, the *R*_0_ was assumed as follows

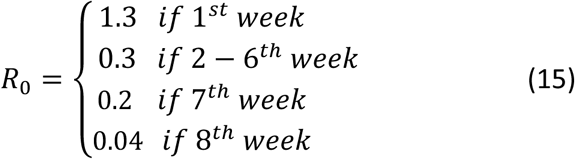
- **“Shelter-in-place” Containment**: It assumes voluntary community-wide home quarantine with only essential services open. In order to capture this intervention, the *R*_0_ was assumed as follows

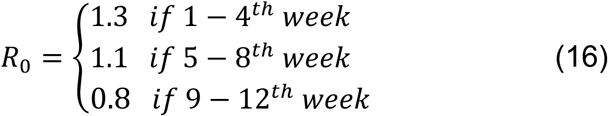
- **“Social Distancing” Order**: It assumes ban on events over 50 people and public advocacy around “social distancing”. In order to capture this intervention, the *R*_0_ was assumed as follows

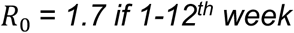

Using these estimates of R_0_ and the formula for R_0_ for the SEIR model with both E and I infectious, it is possible to readily obtain corresponding *β* estimates (transmission rate over time). For intervention *β* will be also piecewise defined function.

**Figure 1.**
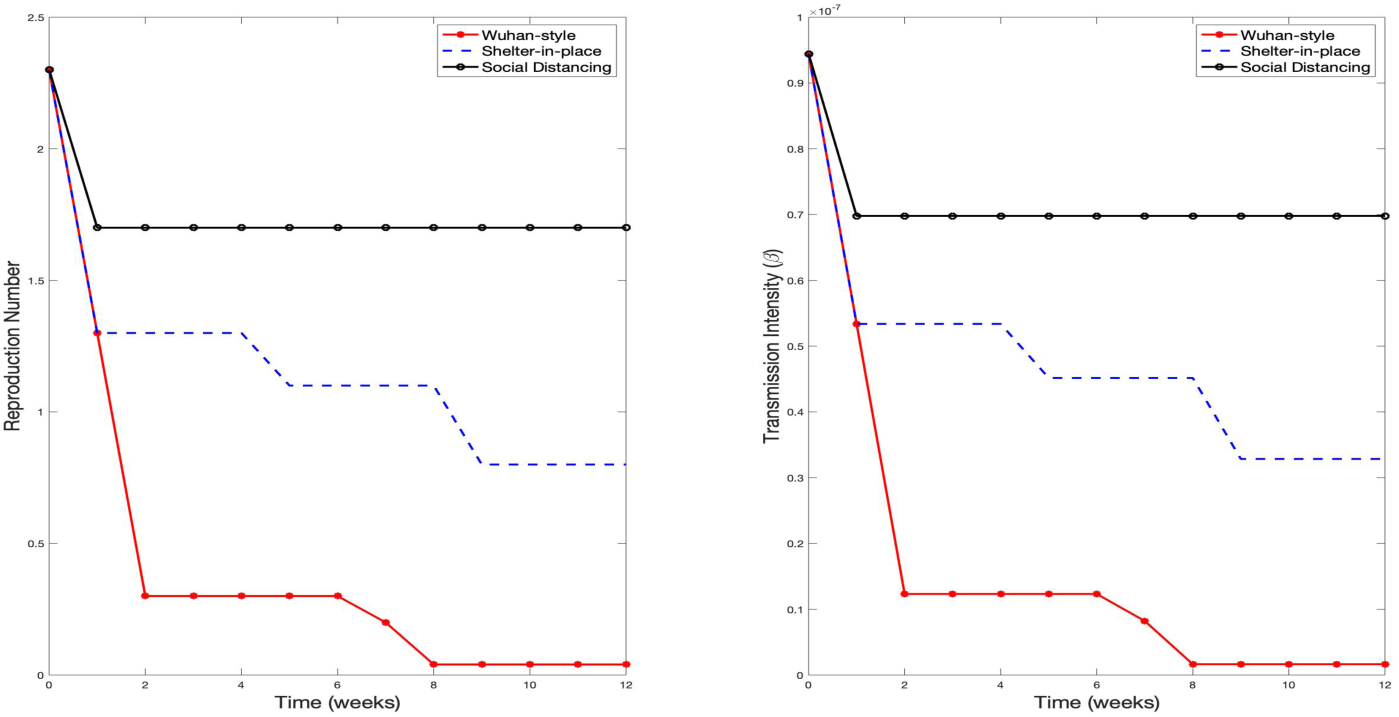
Three different types of transmission metrics corresponding to the three different interventions

## 3. Results

We assume that the initial values for simulations were *I*_0_ *=* 1*,S*_0_ *= N* − *I*_0_ and other state variables zero at the starting time. The parameter estimates of the models were collected in Table 3 and Table 4.

**Table 3.** Date of reporting starts and first exponential day

| Country | Date of reporting starts | Estimated date of first Case (number of days of delay) | Initial reported cases $y(0)$ | $\Lambda$ | R-squared |
| --- | --- | --- | --- | --- | --- |
| South Korea | 02/19/2020 | 02/03/2020 (16 days) | 86 | 0.2861 | 0.89 |
| South Africa | 03/09/2020 | 03/04/2020 (5 days) | 4 | 0.3105 | 0.98 |
| Mexico | 03/11/2020 | 03/02/2020 (9 days) | 8 | 0.2418 | 0.92 |
| Panama | 03/11/2020 | 02/25/2020 (15 days) | 18 | 0.2056 | 0.95 |
| Brazil | 03/10/2020 | 02/24/2020 (15 days) | 36 | 0.2419 | 0.93 |
| Chile | 03/10/2020 | 02/28/2020 (11 days) | 14 | 0.2473 | 0.92 |
| Ecuador | 03/13/2020 | 03/01/2020 (12 days) | 21 | 0.2618 | 0.86 |
| Peru | 03/09/2020 | 02/28/2020 (10 days) | 8 | 0.2231 | 0.89 |
| Argentina | 03/09/2020 | 03/28/2020 (10 days) | 8 | 0.2158 | 0.96 |
| Colombia | 03/09/2020 | 03/02/2020 (7 days) | 5 | 0.2391 | 0.92 |

**Table 4.** SEIR model parameters estimate for *R*_0_ *=* 4.95

| Parameters | Description | Estimated value (units) | Reference |
| --- | --- | --- | --- |
| $\Lambda$ | Growth rate: duration of an epidemic cycle | 0.175 (day<sup>-1</sup>) | Estimated |
| $\lambda$ | Force of infection: rate at which susceptible individuals becoming infected per unit time | $1.54 \times 10^{-4}$ (day<sup>-1</sup>) | Estimated |
| $T$ | Interepidemic period | 5.1 (years) | [7] |
| $A$ | Median Age of infection | 17.7 (years) | [7] |
| $I_t$ | Immunity Threshold: proportion of immune population minimum necessary to extinguish epidemic | 0.8 (proportion population) | [7] |
| $T_s$ | Serial interval | 7.5 (days) | [6] |
| $T_d$ | Doubling time of cases | 7.4 (days) | [6] |
| $D$ | Duration of infectious period | 7 (days) | [7,8,15] |
| $D'$ | Duration of the asymptomatic period | 7 (days) | [7,8,9] |

### Initial Exponential Growth Rate

Exponential growth model was used to estimate initial exponential growth rate in city of Cali. The exponential function (Equation (7) or (8)) is fitted to reported cases from different countries. The early estimated growth rate using data from China is found to around 0.17 (Figure 2) and this is used for further simulation scenarios. The confidence interval is given by 95% CI (0.162, 0.185). The comparison of different exponential growth and estimated start of epidemics in some countries are computed (Table 3).

**Figure 2.**
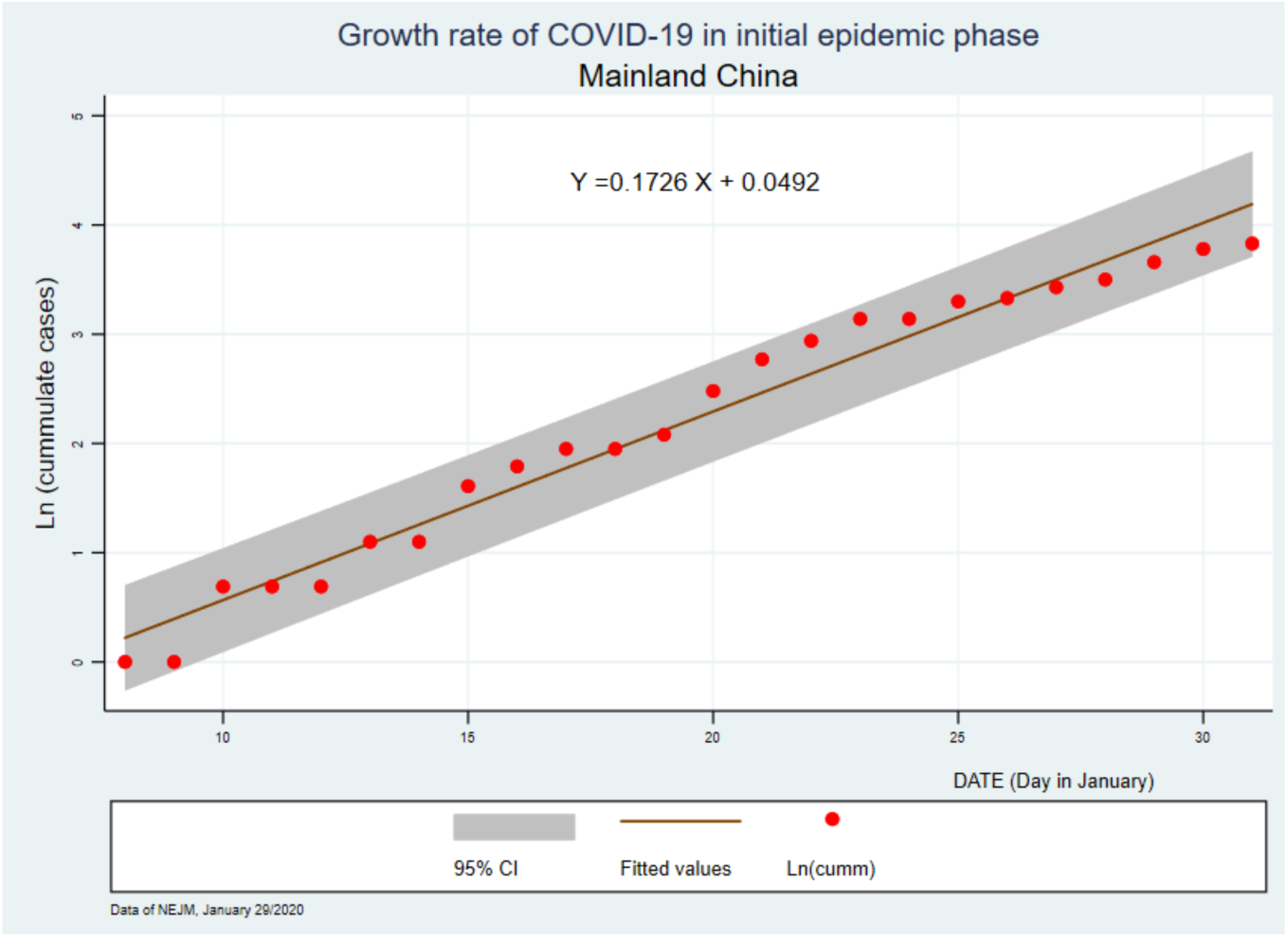
Growth rate of Covid-19 in initial epidemic phase

### Outbreak in City of Cali, Colombia

#### SEIR model with I Infectious

Using literature, we can estimate some of the model parameters. We obtained *T_h_* (the hospitalization rate) around 14%, *L_h_* (the lethality in hospitalized individuals) around 15%, the estimated *T_s_ =* 7.5 days (95% CI, 5.3 to 19) and the estimated *T_d_ =* 7.4 days (95% CI, 4.2 to 14) (6).

It is assumed that the individuals contact randomly and furthermore the controls are not implemented. We carried out simulations with the population size of Cali and it was applied the hospitalization rate (*T_h_*) and the lethality rate (*L_h_*) in hospitalized individuals reported from Wuhan.

Based on the data for confirmed cases from December 08, 2019 through January 21, 2020 from city of Wuhan, China [14], we estimated the growth rate [15] of natural logarithm of the incidence accumulate be 0.175 and the corresponding *R*_0_ was 4.95.

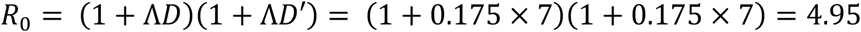

#### Comparison of SIR, SEIR with I only infectious and SEIR with E and I both Infectious models

Various outbreak related-metrics including number of cases and deaths for the city of Cali are given in Table 5 when there are no interventions. If we assume that infections can be generated by asymptomatic then the outbreak will persist for relatively longer time (around 8 months versus 7 months). The total number of hospitalized cases during the outbreak between the three models are presented in Table 6 for 30, 60 and 90 days after start of epidemic, respectively.

**Table 5.** Description of parameters and values for all other models in Table 2.

| Parameter | Description | Value(units) | References |
| --- | --- | --- | --- |
| $\beta$ | Rate at which two specific individuals come into effective contact per unit time | $2.83 \times 10^{-7} \text{ (day}^{-1}\text{)}$ | [5,14] |
| $q$ | Reduce infectivity of E class individual as compared to I individual | 0.5 | Assumed (varied) |
| $l$ | Reduced infectivity of J individual as compared to I individual | 0.4 | Assumed (varied) |
| $\kappa$ | The rate at which individuals in the pre-infectious category become infectious per unit time. $1/D$ | $0.143 \text{ (day}^{-1}\text{)}$ | [7,8,15] |
| $\mu$ | Per capita rate at which asymptomatic get recovered without showing symptoms | $0.2 \text{ (day}^{-1}\text{)}$ | [42] |
| $\eta$ | Recovery rate in treated: the rate at which the infectious are treated and cured | $0.85 \text{ (day}^{-1}\text{)}$ | [9] |
| $\gamma_1$ | Per capita rate at which symptomatic recovered | $0.125 \text{ (day}^{-1}\text{)}$ | [42] |
| $\alpha$ | Recovery rate: The rate at which infectious individuals recover (become immune) per unit time. $1/D'$ | $0.143 \text{ (day}^{-1}\text{)}$ | [7,17] |
| $\delta$ | Per capita disease related mortality rate | $0.025 \text{ (day}^{-1}\text{)}$ | [42] |
| $\sigma$ | Per capita rate at symptomatic individuals are isolated | $0.066 \text{ (day}^{-1}\text{)}$ | [43] |
| $\phi$ | Per capita quarantine rate | $0.20 \text{ (day}^{-1}\text{)}$ | [43] |
| $1/\theta$ | Average time in quarantine | 6 (days) | [43] |
| $\gamma_2$ | Per capita rate of recovery for hospitalized patients | $0.75 \text{ (day}^{-1}\text{)}$ | [42] |
| $D$ | Duration of infectious period | 7 (days) | [7,8,15] |
| $D'$ | Duration of the preinfectious with asymptomatic period | 7 (days) | [7] |

**Table 6.**
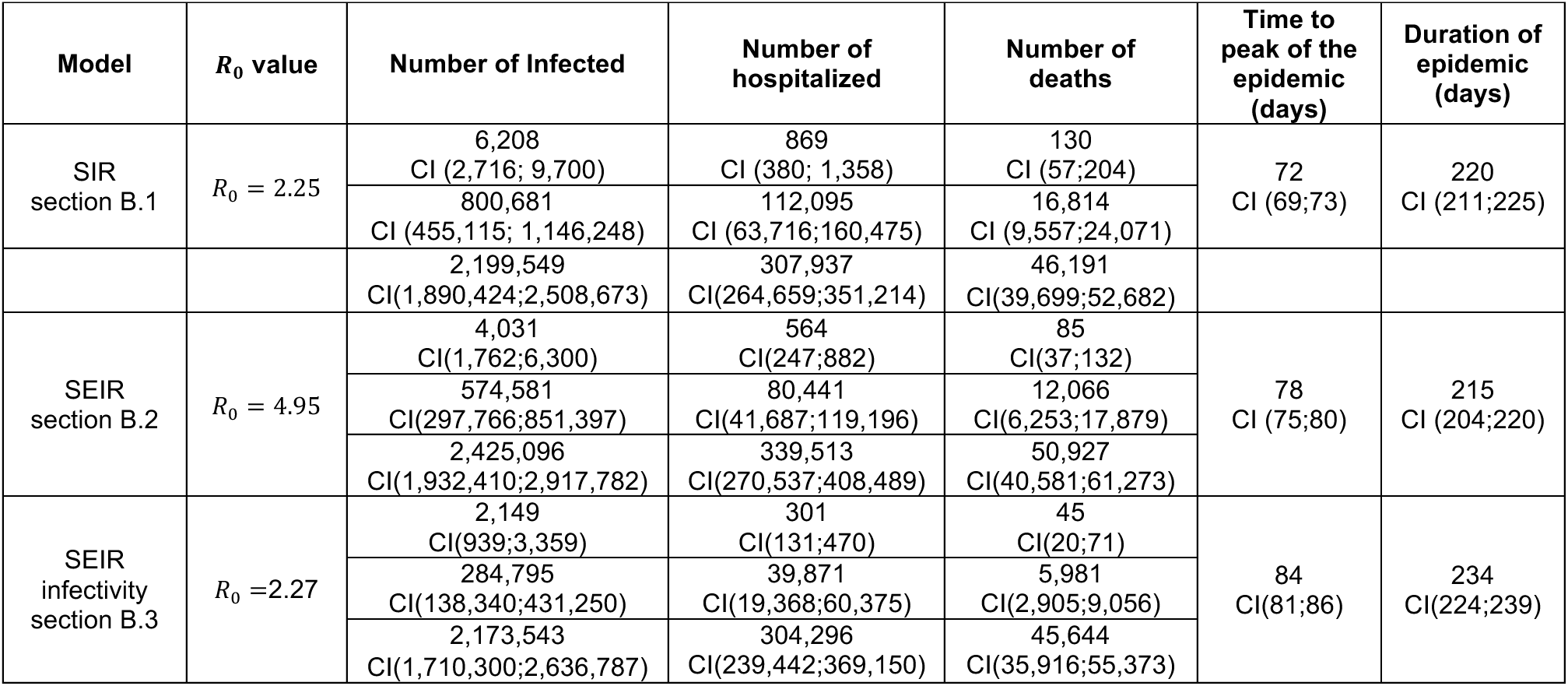
Scenario for different models, SIR, SEIR with only I infectious, and SEIR with *E* and *I* both Infectious. For each model, the first line represents 30 days after start of epidemic, the second line 60 days from beginning and the third line have values after 90 days in epidemic. The number of hospitalized is calculated using the hospitalization rate, *T_h_*, of 14% [15] and number of deaths is calculated using the COVID-19 mortality rate, *L_h_*, of 15% among hospitalized individuals [15].

**Figure 3.**
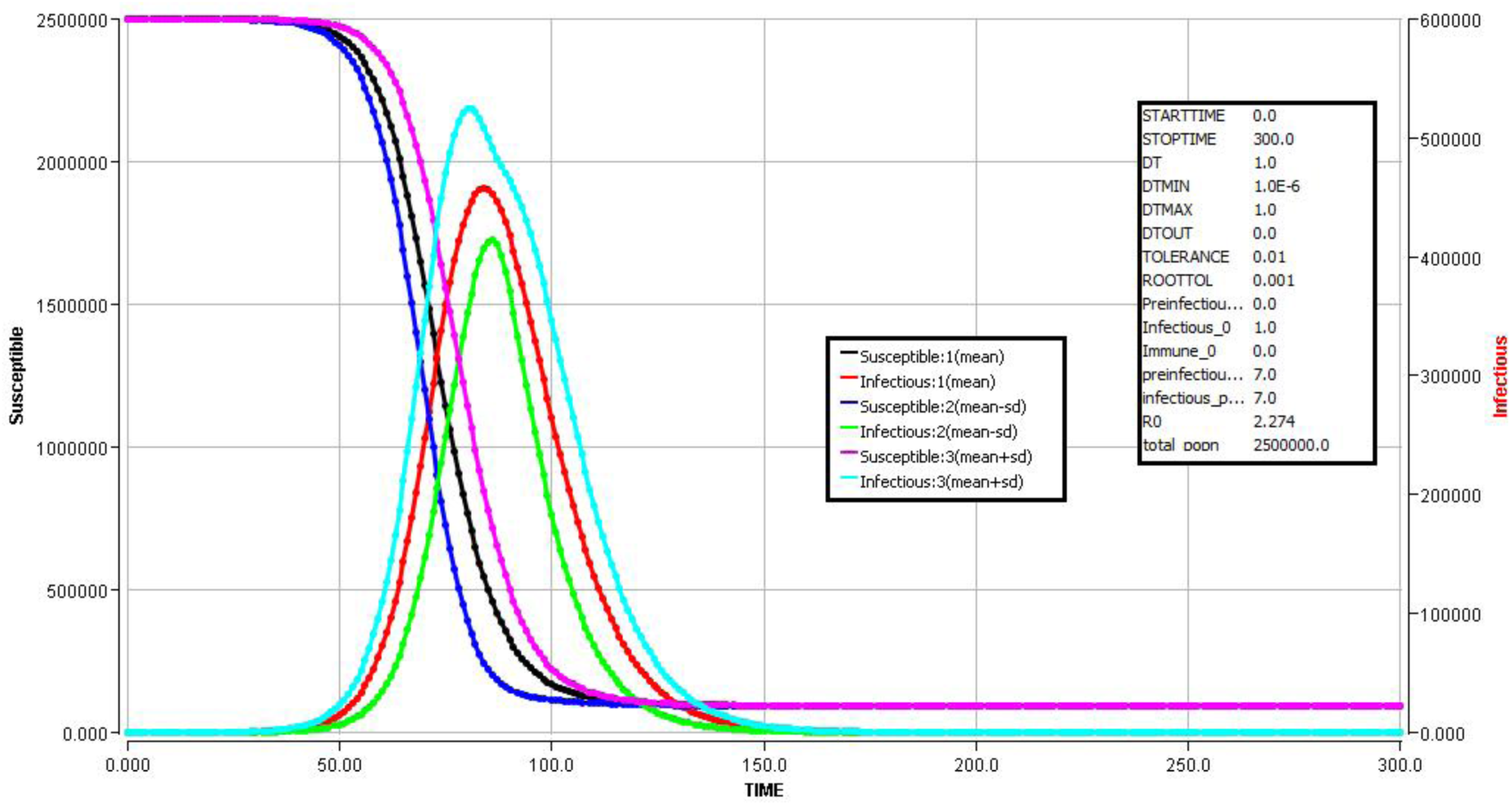
Simulation for SEIR model with *E* and *I* both infectious

#### Comparison of Outbreak burden using SEIR Model with no intervention, only isolation and both quarantine and isolation interventions

We consider SEIR-Q1Q2Q3 model (see Appendix, section B6 for details) under three different scenarios: baseline (no intervention), with only isolation and with both quarantine and isolation (Figure and Table shows the statistics and trends).

**Figure 4.**
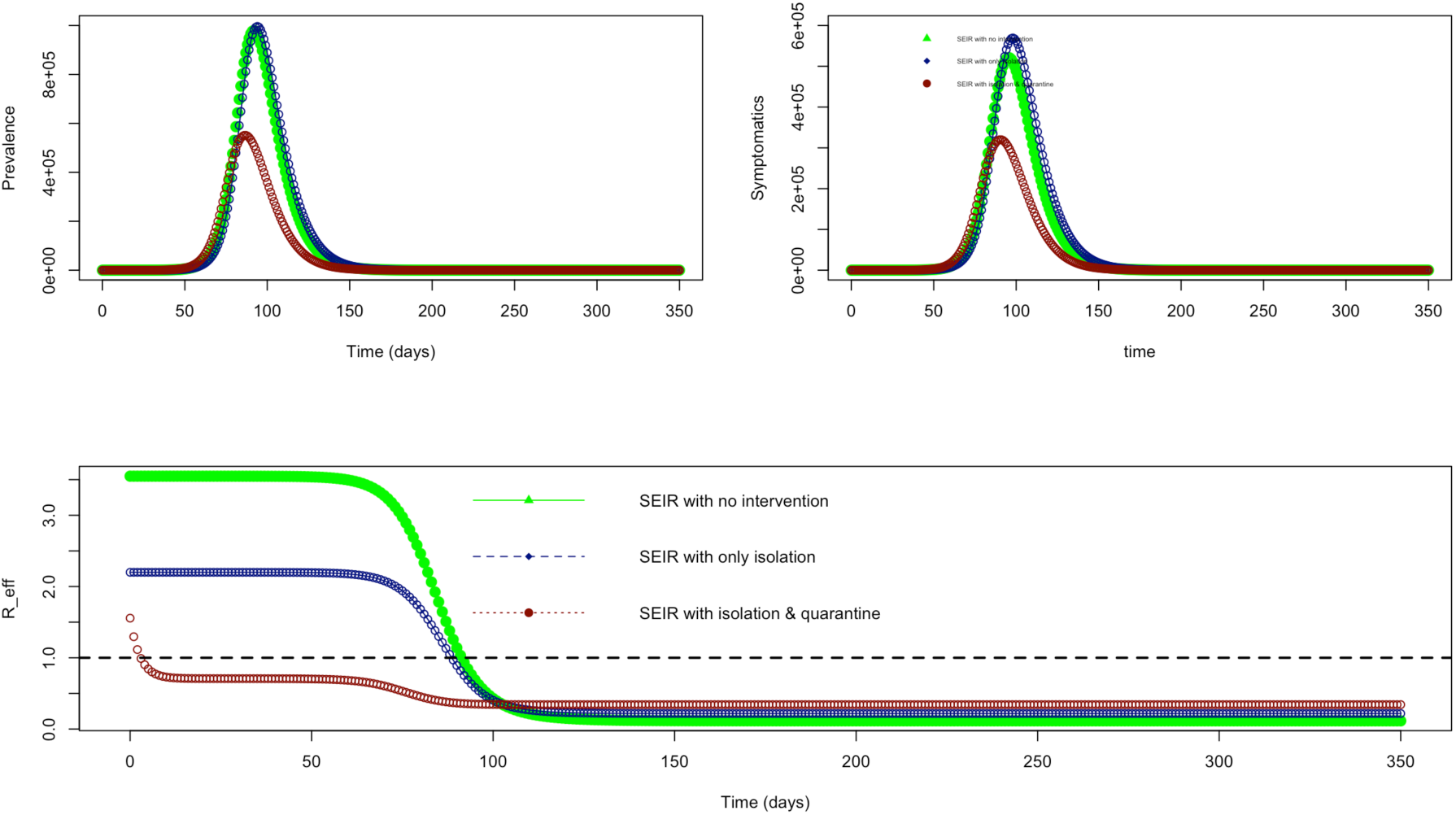
SEIR-Q1Q2Q3 Model results. Showing total cases and effective reproduction number

**Table 7.** SEIR-Q1Q2Q3 Model results

| | $R_0/R_c$ | Beta | End of outbreak (days) | Total Cases | Maximum number of symptomatic on a given day | Number Hospitalized (14%) |
| --- | --- | --- | --- | --- | --- | --- |
| SEIR | 3.54 | 0.3169 | 246 | 2,244,879 | 521,313 | 72,983 |
| SEIR with Q3 | 2.19 | 0.4916 | 209 | 2,090,252 | 95,044 | 13,306 |
| SEIR with Q1, Q2, Q3 | 1.55 | 0.7930 | 160 | 1,196,356 | 25,021 | 3,502 |

**Figure 5.**
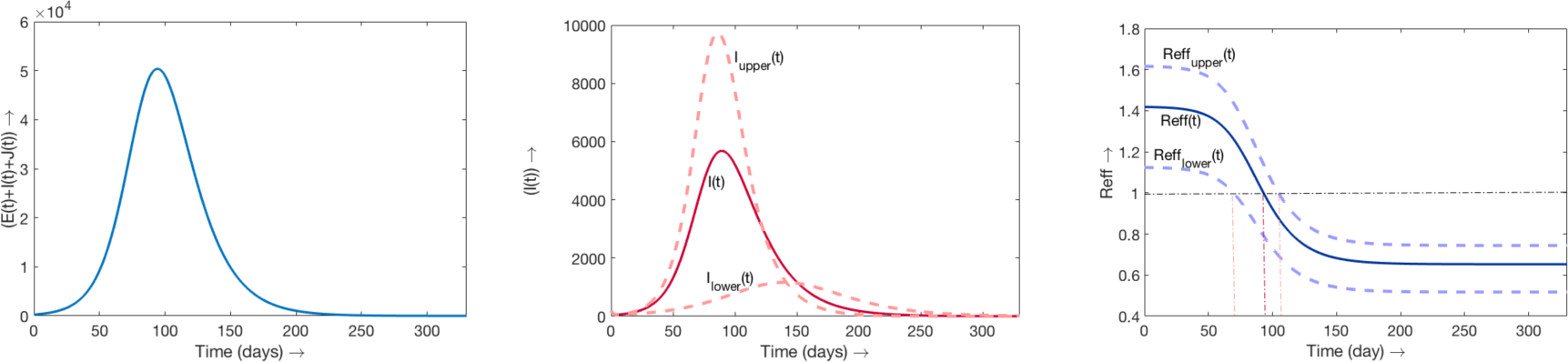
SEIR-J.

### Comparison Between 5 Big Cities of Colombia

#### Comparison of SEIR with E and I both Infectious and quarantine and isolation interventions models for 5 different cities of Colombia

The difference between outbreaks under similar level of quarantine and isolation interventions are shown in Table 8 and Figure 6. Cartagena and Barranquilla will have roughly similar type of an outbreak and Cali and Bogota will have similar. Maximum number of a beds that will be needed on a given day in Bogota and Cartagena will be 3 and 1/2 times, respectively, the corresponding number in Cali. Cali might need up to 3502 beds a day for the COVID-19 patients at the time of the peak of the outbreak, which will last for 7 to 8 months.

**Table 8:** Expected number and 95% confidence interval

| Model | $R_0/R_c$<br>(95% CI) | Time when $R_{eff} < 1$<br>(95% CI) | Duration of an outbreak $[T_{end}]$<br>(95% CI) | Total Number of Cases in Outbreak $[E(T_{end}) + I(T_{end})]$ | Time to Peak of Epidemic (95% CI) | Cases when Peak is reached (95% CI) |
| --- | --- | --- | --- | --- | --- | --- |

|  |  |  |  | (95% CI) |  |  |
| --- | --- | --- | --- | --- | --- | --- |
| SEIR with *I* Infectious | 3.027<br>[2.831;3.231] | 85.36<br>[84.75;85.93] | 163<br>[159;168] | 2185473<br>[2152489;2213376] | 87.7<br>[82.33;93.44] | 187839<br>[214347;187542] |
| SEIR with *E* & *I* infectious | 2.354<br>[2.259;2.463] | 87.51<br>[86.51;88.47] | 169<br>[163;174] | 2021374<br>[1983782;2058871] | 92.54<br>[86.29;100.4] | 165129<br>[157569;188438] |
| SEIR-J | 1.336<br>[1.175;1.520] | 92.86<br>[69.44;104.6] | 302<br>[279;326] | 401937<br>[256819;5318737] | 90.05<br>[87.16;151.6] | 5680<br>[1,128;9,727] |

**Figure 6.**
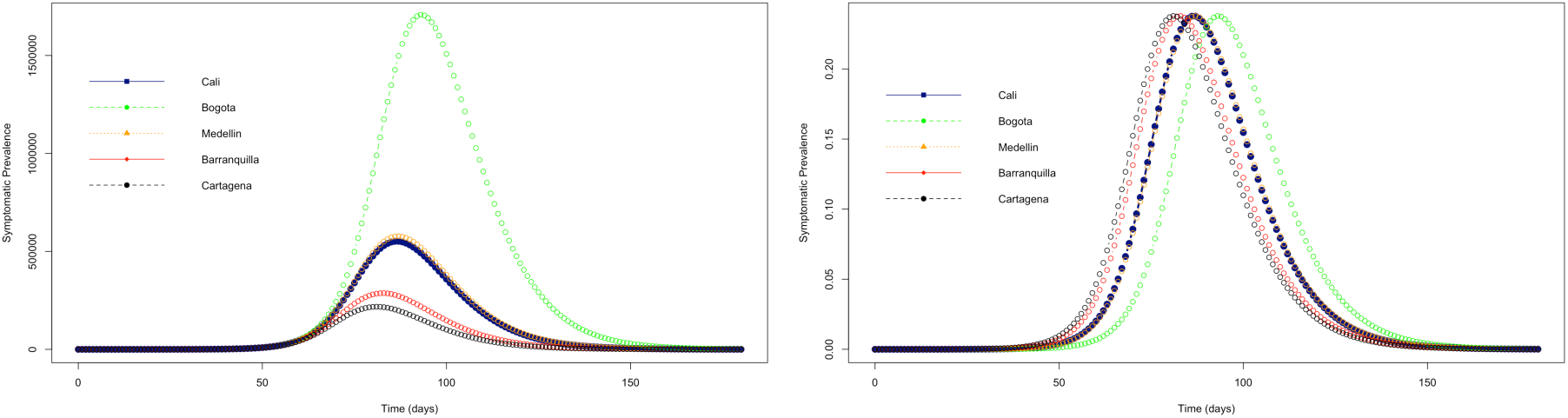
Comparison of outbreaks in 6 major cities of Colombia using SEIR_Q1Q2Q3 model

**Table 9:** SEIR-Q1Q2Q3 Model results for Different Cities. Not all symptomatic will be needing hospitalization as some will recover themselves. In all the cities roughly 52% will be infected either as mild or severe infections during the outbreak

| SEIR with Q1, Q2, Q3 | Population size (N) | End of outbreak (days) | Total Cases [Symptomatic + Asymptomatic] | Maximum number of symptomatic on a day | Max number hospitalized in a day (14%) |
| --- | --- | --- | --- | --- | --- |
| Cali | 2.3M | 236 | 1196356 | ~25021 | ~3502 |
| Bogota | 7.2M | 254 | 3703776 | ~77423 | ~10839 |
| Medellin | 2.4M | 237 | 1251704 | ~26178 | ~3664 |
| Barranquilla | 1.2M | 226 | 621984 | ~13006 | ~1820 |
| Cartagena | 0.9M | 221 | 471673 | ~9863 | ~1380 |

## 4. Discussion

The estimates of *R*_0_ considered here might be slightly higher than those published by other authors, from Asia and Europe [21,32,34] but it is noteworthy that during the SARS epidemic in 2002 the estimates of the average R_0_ varied between 2.24 (95 % CI: 1.96–2.55) and 3.58 (95% CI: 2.89–4.39) [4] and 8,000 cases were reported while for the new SARS-Cov-2 which is a similar coronavirus, originated in the same country the estimates of *R*_0_ are similar or even lower than 2.2 (95% CI, 1.4 to 3.9) [14], and 80,000 cases have already been reported in the first 11 weeks of the epidemic.

We have observed the application of different techniques to calculate R_0_, that could explain the different scenarios of a potential outbreak in Colombia. While Li et al. (14) used the duration of the serial interval, whose result is similar to ours when we use the same technique, our highest estimate was based on the growth rate and the duration of similar pre-symptomatic and infectious periods.

We estimated that in the city of Cali the outbreak under current intervention of isolation and quarantine will last for 5-6 months and will need around 3500 beds on a given during the peak of the outbreak. Outbreak in Cali will be similar to outbreak in Medellin but at least 1/3 times less cases will be observed as compared to the outbreak in Bogota. The cases in the current outbreak in Cali was reported about a week late.

## 5. Conclusions

COVID-19 is a highly transmissible virus with the capacity to produce outbreaks and with high repercussions on lethality in the vulnerable (individuals with pre-health conditions or elderly) population and with comorbidities.

Mathematical models and simulation applied to data ongoing epidemics allow anticipation in the phase of preparing mitigation plans.

Based on the estimates, the distribution of hospital beds and intensive care units in the city available for response to an eventual emergency have been planned. Also, the inclusion of 500,000 Euros in municipal budget for mitigating the epidemic if it reaches.

## Data Availability

We used data obtained from different documents already published online.

## Appendix

### A. Some other figures

**Figure 7.**
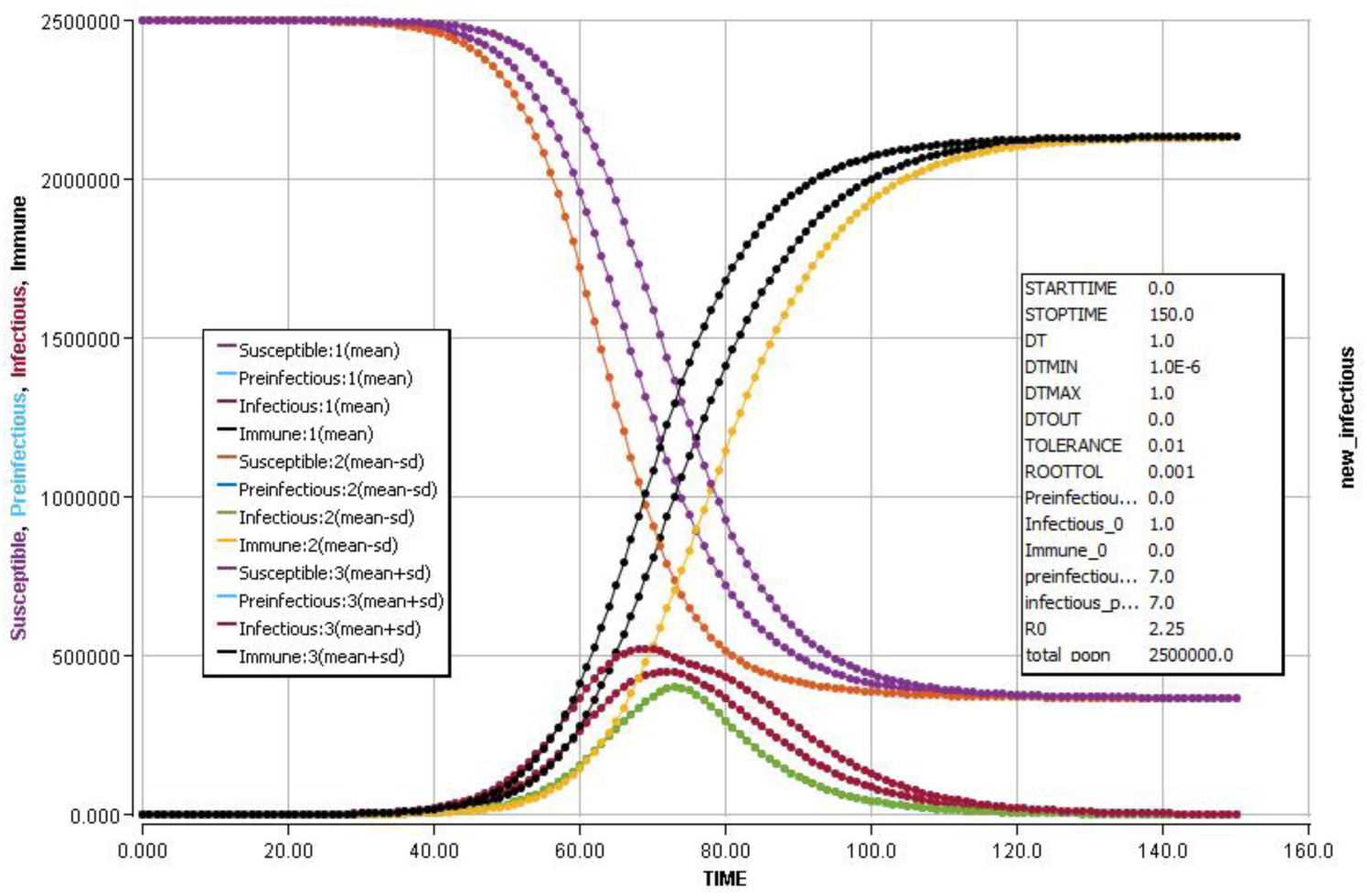
Simulation for SIR model

**Figure 8.**
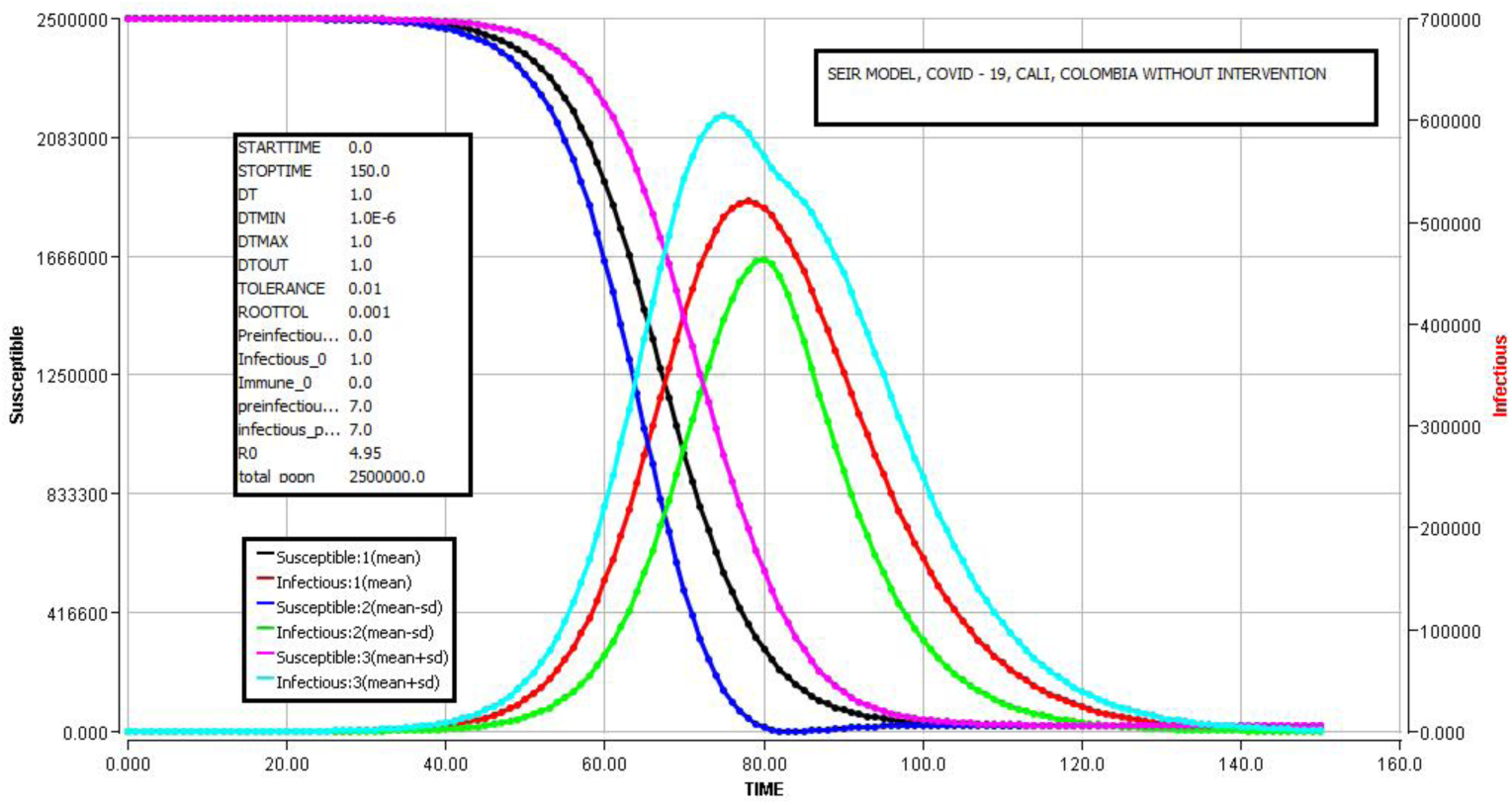
Simulation for SEIR model with only *I* infectious

**Figure 9.**
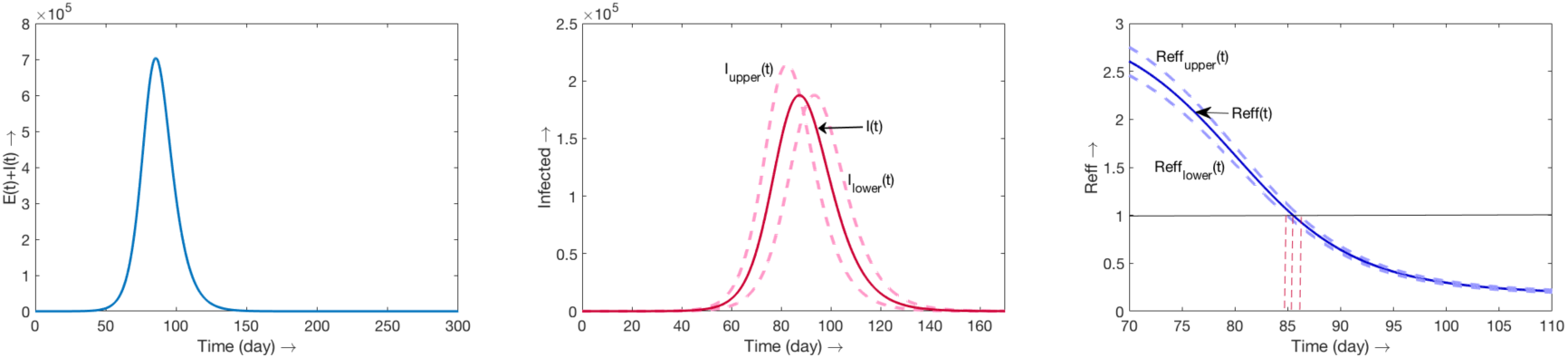
SEIR with only I infectious

**Figure 10.**
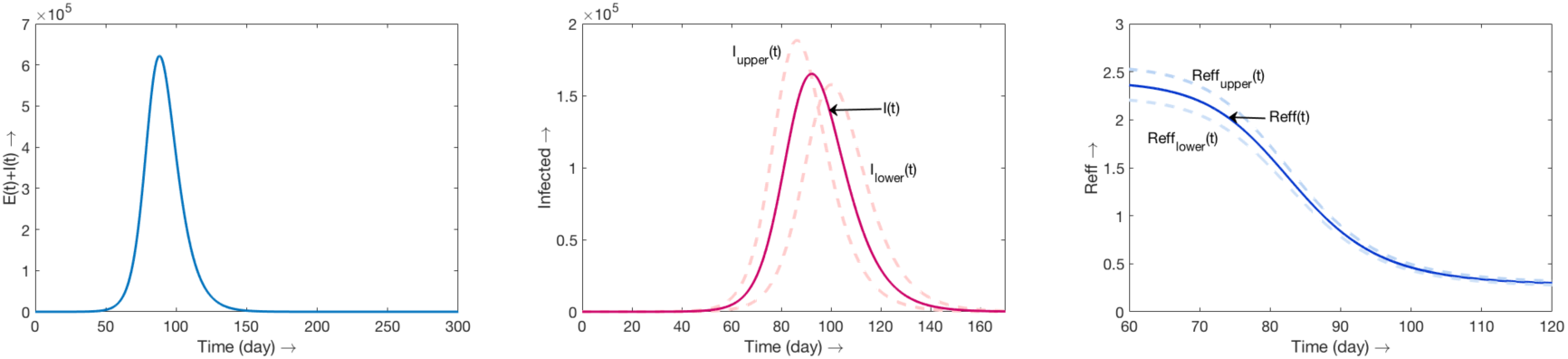
SEIR with E and I infectious

**Figure 11.**
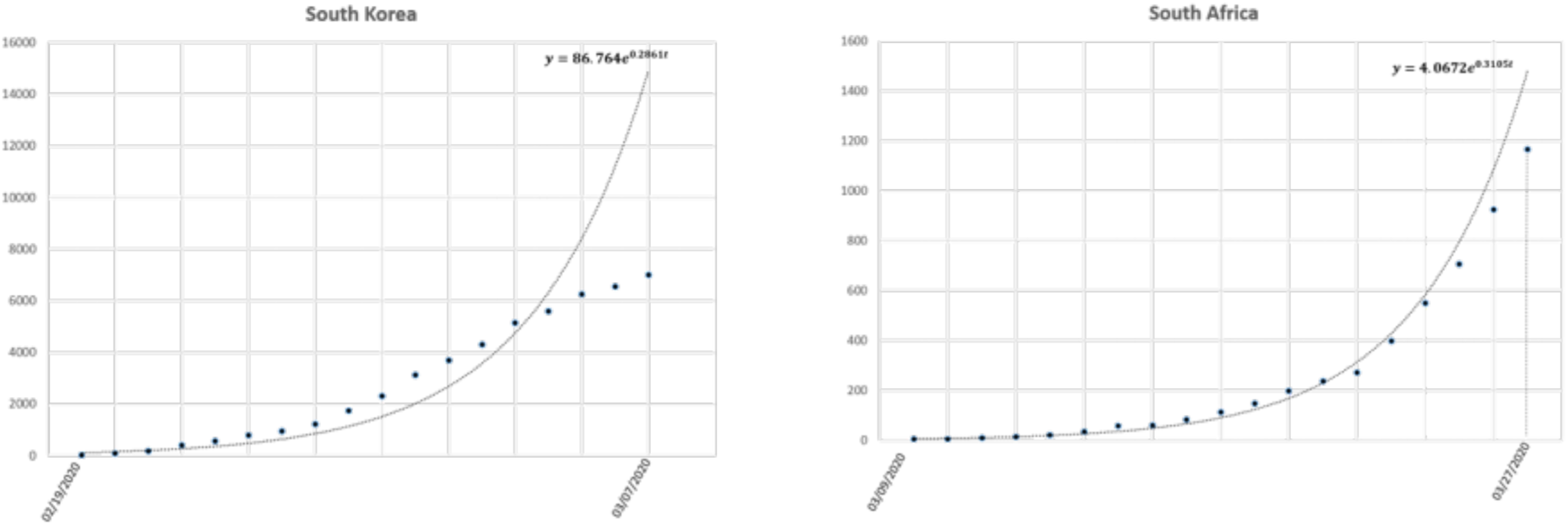
Exponential fitting for data from South Korea and South Africa

**Figure 12.**
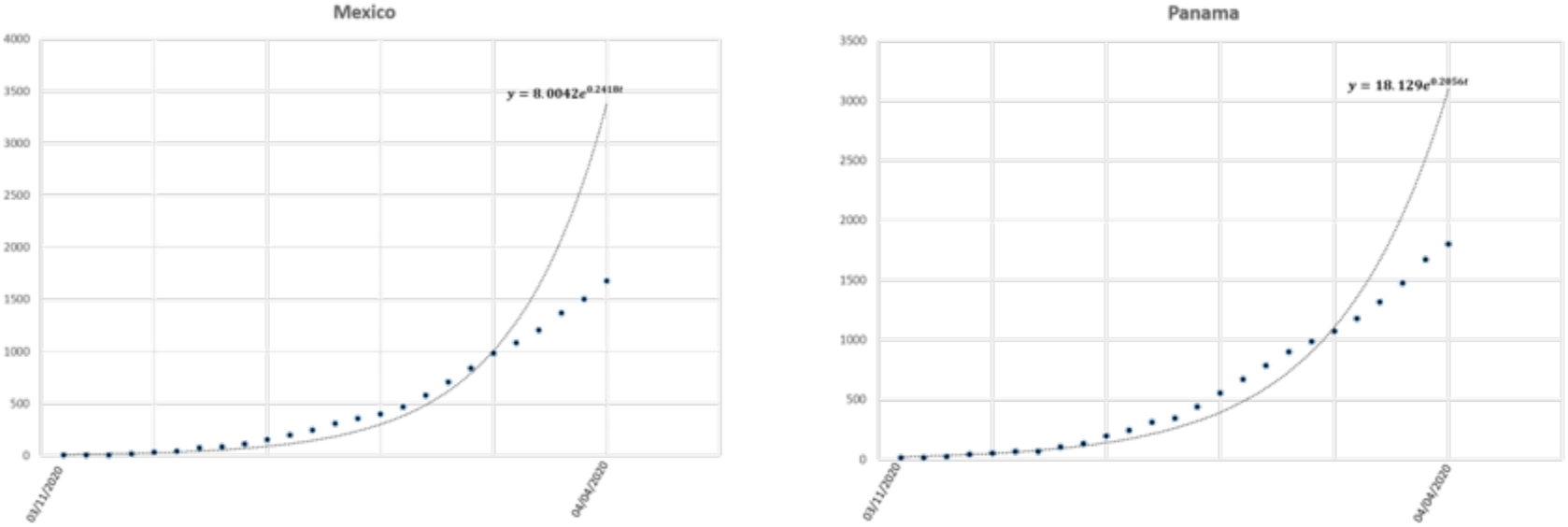
Exponential fitting for data from Mexico and Panama

**Figure 13.**
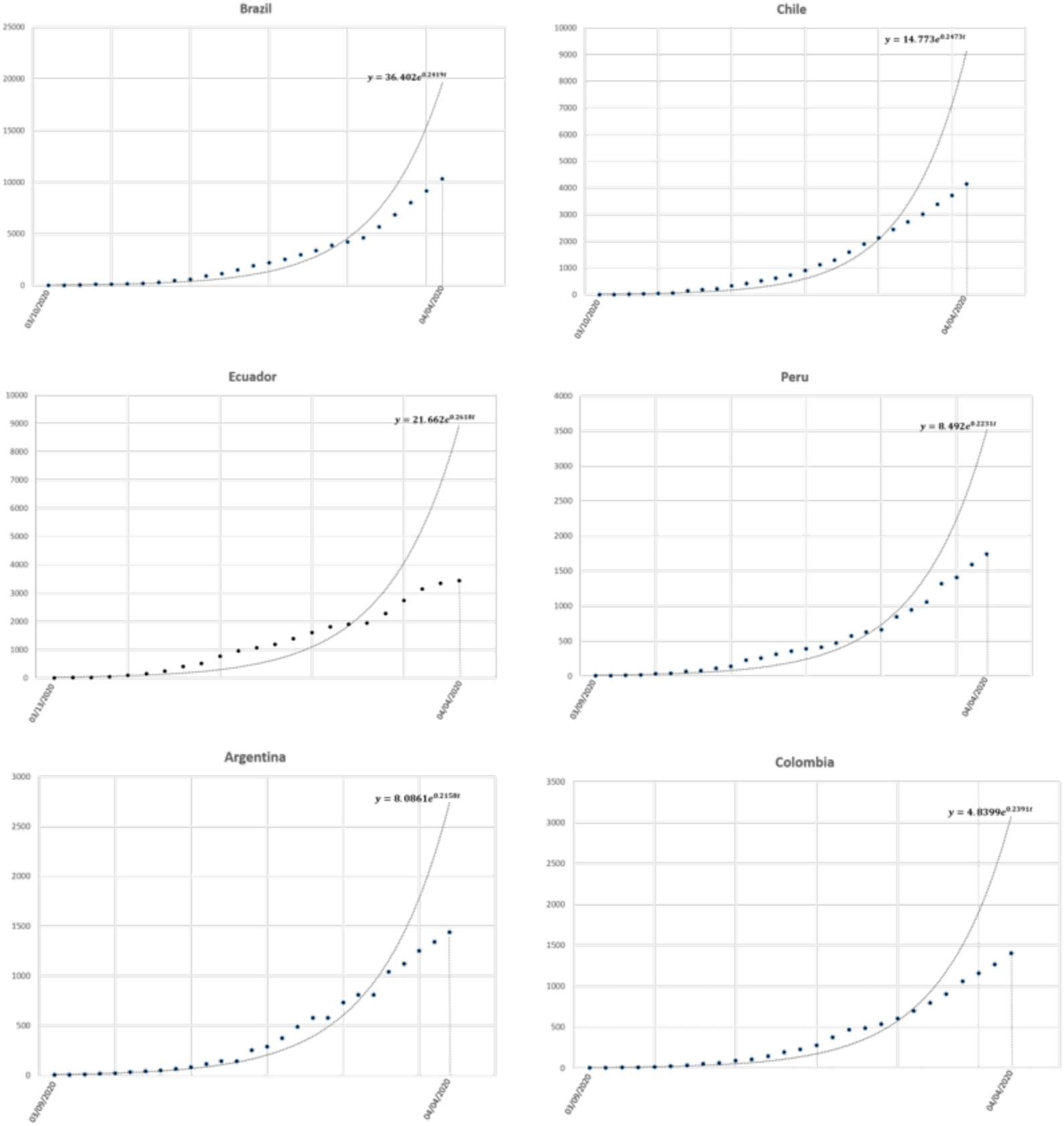
Exponential fitting for data from six South America countries

### B. Models

#### B.1. SIR Model

We start with a simple SIR epidemic model

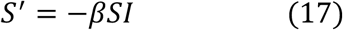

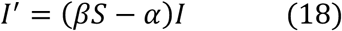

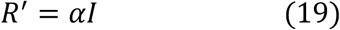

with initial conditions *S*(0) = *S*_0_*, I*(0) = *I*_0_, *S*_0_ +*I*_0_ = *N*. It is known that the basic reproduction number for this model is 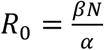 and in terms of the initial exponential growth rate Λ

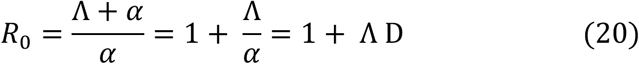

For this simple SIR model, if t is small, *S* ≈ *N*, and the equation for *I* is approximately

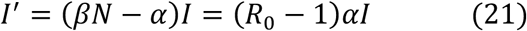

and grow exponentially with growth rate (*R*_0_ − 1)*α*. The exponential growth rate A can be measured and then we have an estimate

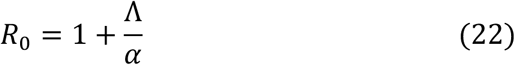

More complicated models are approximated for small *t* by linear systems, whose solutions have an exponential growth rate given by the largest eigenvalue of the coefficient matrix. Thus for the SEIR model, the initial exponential growth rate Λ < *α*(*R*_0_ − 1) is the (unique if *R*_0_ *>* 1) largest positive eigenvalue of Jacobian at disease free equilibrium.

The final size relation in terms of the initial exponential growth rate is

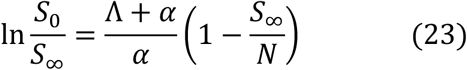

#### B.2. SEIR Model

Now we add an exposed class and we have the following model

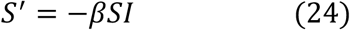

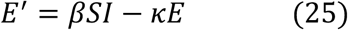

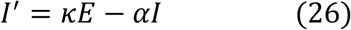

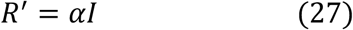

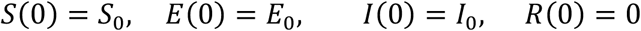

The basic reproduction number for this model is 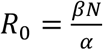 and in terms of the initial exponential growth rate Λ takes the form

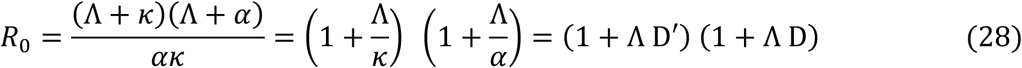

For this SEIR model, the initial exponential growth rate *Λ* is the largest positive eigenvalue of

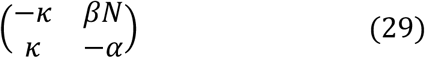

The final size relation for this model in terms of the initial exponential growth rate is

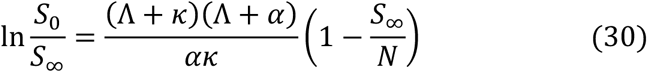

If we estimate *R*_0_ from other formulas obtain

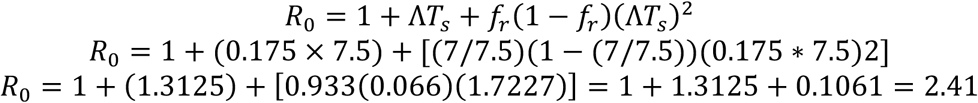

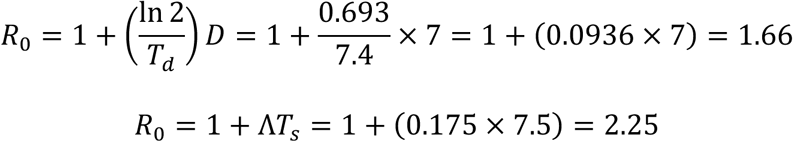

We have estimated the expected number of 255 infected, 9 hospitalized and 1 death, during the first month of the epidemic if one infected individual with SARS-COV-2 enters Cali and no control would be taken.

#### B.3. SEIR Model with infectivity in exposed stage

Now we consider the last model with infectivity in exposed stage

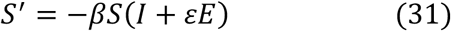

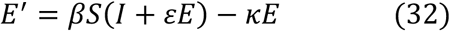

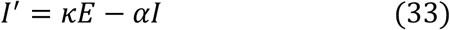

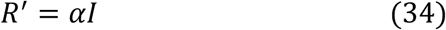

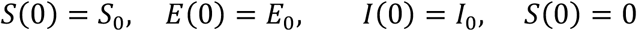

In this case the basic reproduction number is 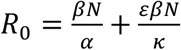 and in terms of the initial exponential growth rate Λ takes the form

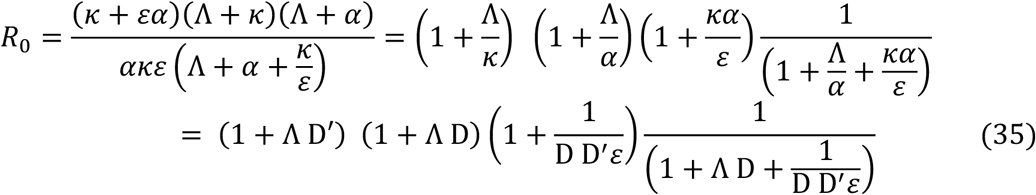

The final size relation in terms of the initial exponential growth rate is

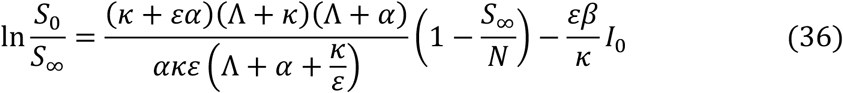

#### B.4. A model with treatment

Now we add treatment to the basic model at a rate *γ*. We consider the following assumptions

- Treatment moves infectives to a class *T* with infectivity decreased by a factor *δ* and with a recovery rate *η*).
- Treatment continues so long as an individual remains infective.
- Treatment is beneficial, *η > δα*.

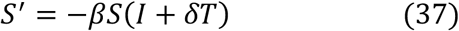

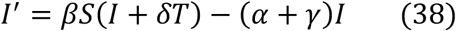

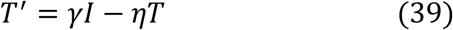

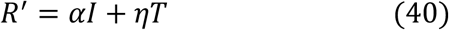

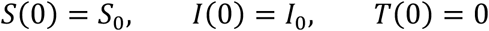

Integration of the first equation, sum of the of the first two equations and the third equation gives

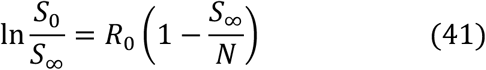

where the basic reproduction number is 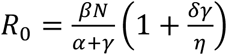 and in terms of the initial exponential growth rate Λ takes the form

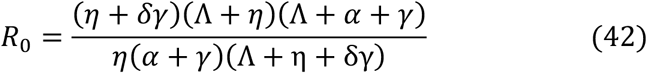

The final size relation in terms of the initial exponential growth rate is

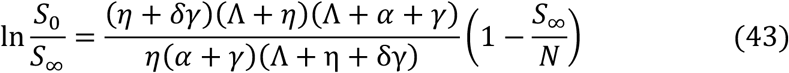

#### B.5 Age of infection model

The age of infection plays a crucial role to determine the rate of disease propagation due to infecting the susceptible individuals. Let *S*(*t*) and *φ*(*t*) denote the number of susceptibles and total infectivity at time *t*. The total infectivity is measured as the average of product of the number of infected individuals and the mean infectivity of the infected individuals at their age of infection *τ* where *τ* ∈ [0, *t*]. Denoting the mean infectivity at time *τ* by *A*(*τ*), we can write the epidemic model with age of infection as follows

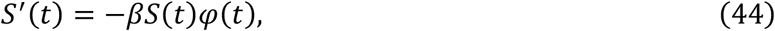

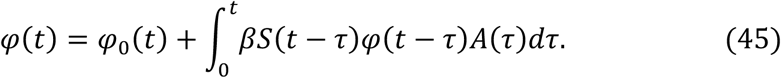

Then the basic reproduction number is defined as

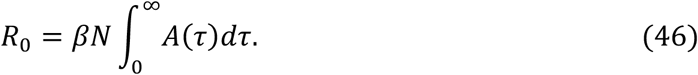

From (44) and (45) we can write

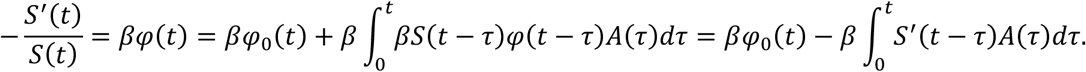

Integrating above expression from 0 to ∞ we get,

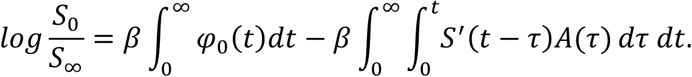

Interchanging the order of integration in the second term at right hand side, we find

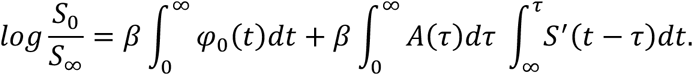

Finally using the straight forward result 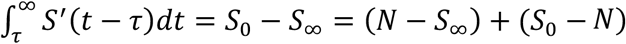, we get the final size relation as

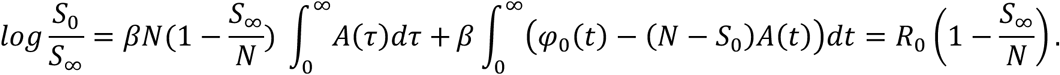

Note that the initial measure of total infectivity at the age of infection *t* is given by *φ*_0_(*t*) *=* (*N* − *S*_0_)*A*(*t*).

This part was done following the book by Brauer, Castillo-Chavez and Feng [41].

The age of infection model is given by

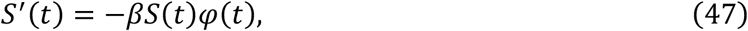

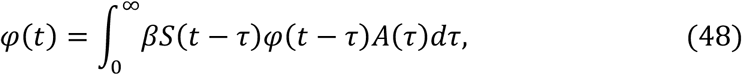

And the basic reproduction number is defined as in equation (46). Now, we linearizing the system (47) – (48) around (*S,φ*) *=* (*N*,0) and find,

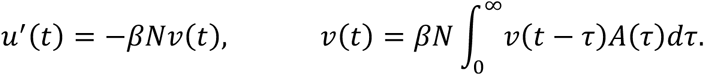

Assuming solution of the above system of equations of the form *u*(*t*) *= e^λt^, v*(*t*) *= e^λt^*, we find one negative eigenvalue *λ =* −*βN* and the other eigenvalue can be obtained by solving

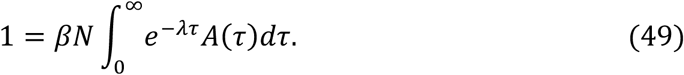

Eliminating *βN* between above equation and (46), we find the basic reproduction number in terms of initial growth rate as follows,

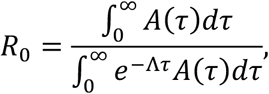

where Λ is a positive eigenvalue satisfying the relation (49).

To derive the final size relation, we can divide equation (47) by *S*(*t*) and then integrating we get,

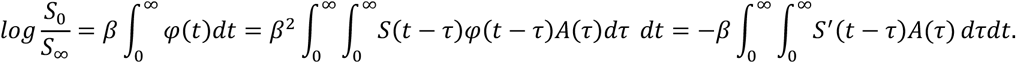

Interchanging the order of integration and using the fact that *S*(−*τ*)= *N*, we can write

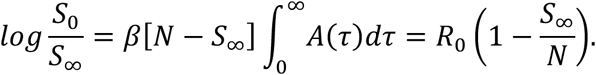

#### B.6. First model with quarantine

Now we consider a model for the transmission of an emerging infectious disease like, the population in the dynamic model is divided into seven classes: Susceptible (*S*), Exposed (*E*), Infectious (*I*), Recovered (*R*), Quarantined (*Q*_1_*,Q*_2_), and Isolated (*Q*_3_). The flow chart for our model is

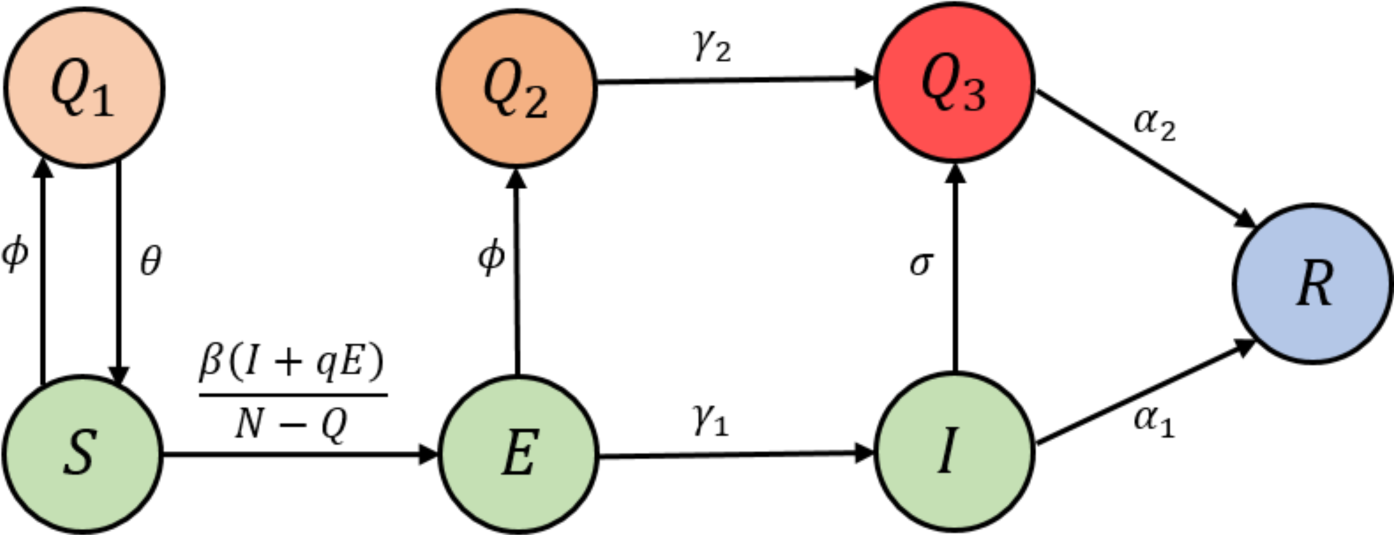

Then the corresponding system of equations is

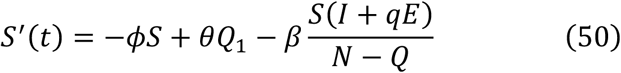

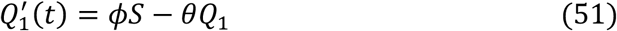

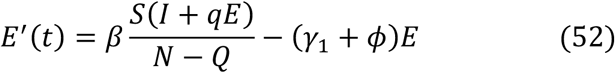

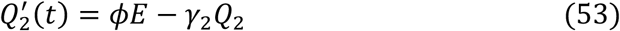

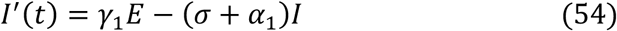

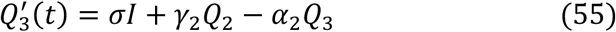

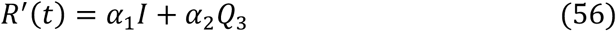

The control reproduction number for this model is

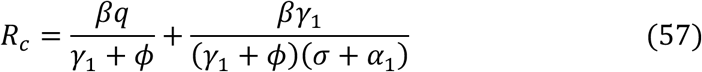

Suppose if 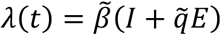 in our system then the control reproduction number becomes

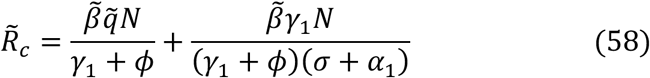

The control reproduction number (*58*) in terms of the initial exponential growth rate Λ is

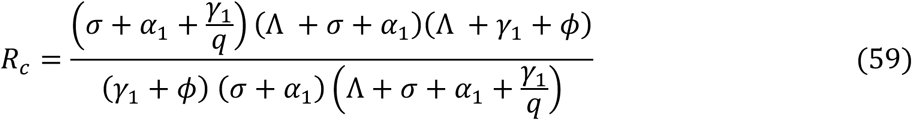

#### B.7. Model with hospitalization and exposed infectiousness

We consider a model for the transmission of an emerging epidemic disease consisting with susceptible (*S*), exposed (*E*), infected (*I*), quarantined (*J*), and recovered (*R*), class as follows,

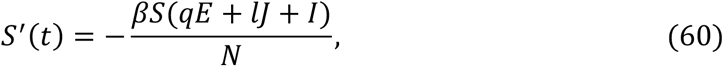

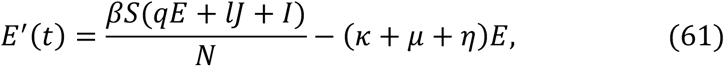

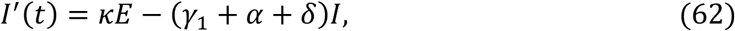

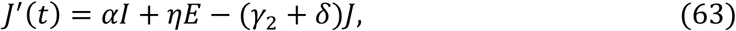

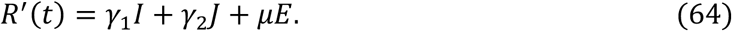

The control reproduction number for this model is

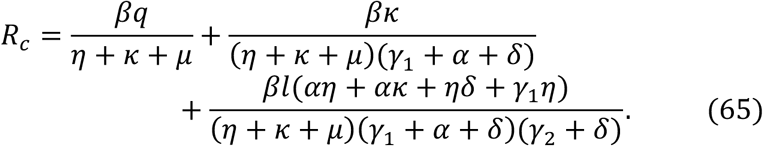

This can be calculated following the approach outlined by Watmogh and P. Ven Den Driesse in *Math. Biosci*. With the help of following two relevant matrices,

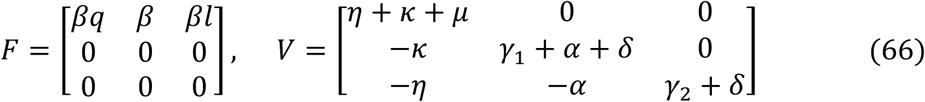

The control reproduction number is the largest eigenvalue of the matrix *FV*^−1^. In order to find the final size relation, integrating both sides of equation (60), we find

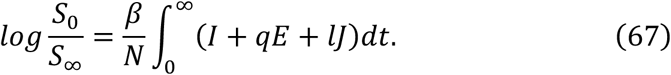

Integrating equation (63) and using the fact that *I*_0_ = 0, *I_∞_ =* 0, we get

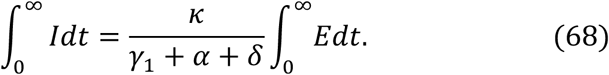

Next we integrate (64), use *J*_0_ = 0, *J_∞_ =* 0, and with the help of (68), we find

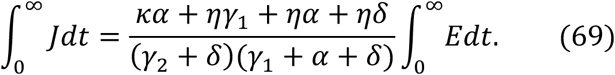

Adding the equations (61) and (62) and then integrating, we find

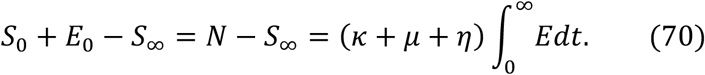

Finally, substituting the results (68) and (69) in (67) and then using (70), after some algebraic calculations, we find

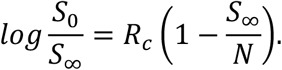

Part of the characteristic equation of the Jacobian matrix for the model (60) – (64) evaluated at disease free equilibrium point which is involved with the determination of initial exponential growth rate Λ takes the form

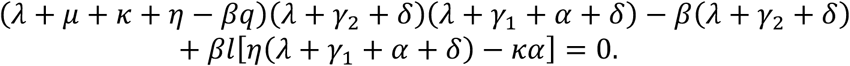

The initial exponential growth rate Λ satisfies above equation and hence we find

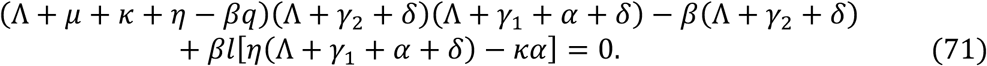

From the controlled basic reproduction number we can write 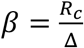, where

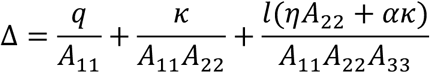

with *A*_11_ *= η + κ + μ, A*_22_ *=γ*_1_ *+ α + δ, A*_33_ *=γ_2_* + *δ*. Using these expressions now we can write (71) as follows,

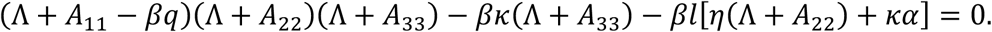

Substituting 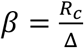 in above equation and then after some algebraic calculation, we find

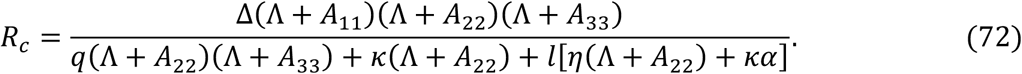

